# Where Are Rabies Cases Lost Along the Surveillance Pathway? A Comparative Evaluation of Participatory and Passive Rabies Surveillance in Urban Lilongwe, Malawi

**DOI:** 10.1101/2022.12.21.22283632

**Authors:** Precious Innocent Mastala, Patrick Ken Kalonde, Edson Chiweta, Tinotenda Razemba, Madeline Nyamwanza, Gracious Chimbalanga, Khumbo Matthew Phiri, Dalitso Lambulira, Precious Dzimbiri, Loveness Kamanga, Mwayi Msowoya, Melaku Tefera, Thoko Flav Kapalamula

## Abstract

**Background:** Rabies remains under-ascertained in many endemic settings, limiting effective surveillance and timely public health interventions, particularly in low-resource countries. Participatory surveillance has the potential to improve case detection, but evidence of its effect on surveillance attrition, case ascertainment, and public health decision-making remains limited. We evaluated a participatory rabies surveillance programme implemented in urban Lilongwe, Malawi, during a six-month pilot (January–June 2020).

**Methods:** Programme records from the pilot were retrospectively analysed. Surveillance progression was tracked from reporting through traceability assessment, investigation, and final case classification. Case retention, case ascertainment, and diagnostic yield were compared between participatory and passive surveillance pathways. Historical rabies diagnostic records from corresponding January–June periods during 2015–2019 served as a comparator.

**Results:** Of 610 suspected rabies reports, 371 (60.8%) were lost before traceability could be established, and a further 89 (14.6%) did not progress from traceability assessment to investigation and final classification. Overall, only 150 reports (24.6%) completed the surveillance pathway. Participatory surveillance retained all reports through classification compared with 104 of 564 (18.4%) passive reports (p < 0.001). Among classified events, confirmed or probable rabies accounted for 13 of 46 (28.3%) participatory reports compared with 7 of 104 (6.7%) passive reports (OR 5.39, 95% CI 1.82–17.39; p = 0.001). Participatory surveillance identified 13 of 20 (65.0%) confirmed or probable rabies cases, including all nine laboratory-confirmed cases detected during the study period. Compared with historical routine surveillance, participatory surveillance was associated with a 2.4-fold increase in laboratory-confirmed rabies detection and a 4.8-fold increase in overall case ascertainment. Among investigated bite exposures, 136 of 141 (96.5%) initiated post-exposure prophylaxis (PEP), although only 16 (11.8%) were linked to confirmed or probable rabies exposures.

**Conclusion:** Rabies cases were lost predominantly between initial reporting and traceability assessment, with further attrition before investigation and final classification. Participatory surveillance reduced surveillance losses, improved case ascertainment, and generated more timely animal health evidence to support One Health public health decision-making, including more appropriate PEP use.

## 1.0 Introduction

Accurate surveillance remains a central challenge for rabies control in endemic settings. Although global estimates suggest that tens of thousands of people die from rabies each year [1], reported cases consistently underestimate the true burden of disease because surveillance systems frequently fail to detect, investigate, and confirm transmission events [1,2]. This under-ascertainment is particularly pronounced in low-resource settings, where laboratory capacity is limited [3–6], and surveillance relies heavily on passive reporting. A useful way to conceptualise this limitation is through a surveillance cascade comprising sequential stages of case recognition, reporting, investigation, sample submission, and laboratory confirmation [7], each of which must function effectively for cases to be ascertained. Failure at any stage results in case attrition, with implications for burden estimation, outbreak detection, and timely intervention, including post-exposure prophylaxis (PEP).

Participatory surveillance, defined as a bidirectional process of collecting and sharing information to support timely action through the active involvement of the target population [8,9], has emerged as an approach to overcome these limitations by expanding disease reporting beyond conventional surveillance systems [10–12]. Unlike passive surveillance, which relies on reporting through routine health or veterinary systems [13], participatory surveillance engages communities and other stakeholders in generating and sharing surveillance information through community-based, integrated, or digital mechanisms [14,15]. In rabies, this broad concept includes approaches such as community-based rabies surveillance (CBRS), integrated bite case management (IBCM) [16], and related community-engaged models that differ operationally but share the goal of strengthening case detection and response.

Rather than simply increasing case counts, participatory surveillance may improve case ascertainment by acting at multiple points within the surveillance cascade. Community engagement and more active reporting mechanisms may support earlier recognition of suspect cases and reduce losses before investigation and final classification. Participatory approaches have improved outbreak detection [17], reporting timeliness [18,19] across a range of human and animal diseases. Evidence from multiple rabies-endemic settings has shown that participatory and community-engaged surveillance approaches can improve reporting and increase identification of animal rabies cases relative to routine surveillance systems [12,16,20–22]. However, existing evaluations have largely focused on overall increases in reporting or case counts, providing limited insight into where along the surveillance pathway these gains can occur.

Understanding where cases are retained or lost is critical for identifying surveillance bottlenecks and determining how participatory approaches contribute to case ascertainment relative to conventional passive systems and to downstream public health decision-making, including PEP administration. Using data from a CBRS programme implemented in urban Lilongwe, Malawi, this study evaluates the contribution of participatory surveillance to rabies case ascertainment relative to routine passive surveillance. Progression across the surveillance cascade, from initial reporting to final case classification, is compared between participatory and passive pathways, alongside patterns of PEP administration among reported exposure events. Quantifying attrition across surveillance stages provides insight into how participatory approaches influence case detection and progression within rabies surveillance systems.

## 2.0 Materials and Methods

### 2.1. Study Area

The study was conducted in urban Lilongwe, the capital city of Malawi (Figure 1), which, at the time of the 2018 Population and Housing Census, had an estimated population of 989,318 residents living in 230,265 households across 58 residential areas [23]. Lilongwe is characterised by rapid urbanisation, heterogeneous settlement structures, and uneven access to services, factors that may influence patterns of human-animal interaction relevant to rabies transmission and control.

**Figure 1.**
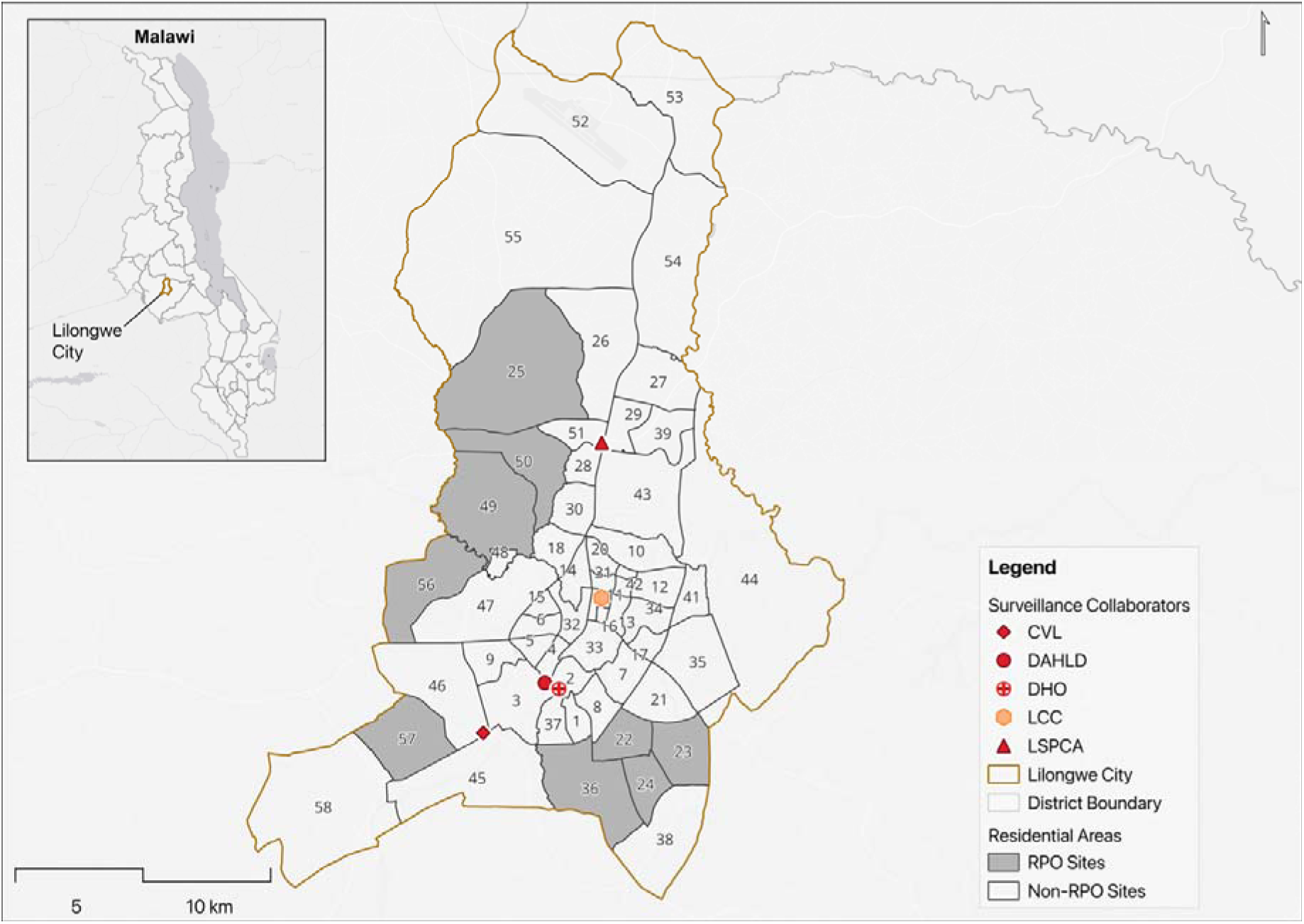
Map of urban Lilongwe showing locations of rabies surveillance partner institutions and residential areas prioritised for deployment of Rabies Project Officers (RPOs). Numbers correspond to residential area identifiers. Abbreviations: LSPCA, Lilongwe Society for the Protection and Care of Animals; DHO, District Health Office; DAHLD, Department of Animal Health and Livestock Development; CVL, Central Veterinary Laboratory; LCC, Lilongwe City Council; RPO, Rabies Project Officer.

Rabies remains endemic in Malawi and continues to represent an important public health concern [24–28]. Approximately 500 human rabies deaths are estimated to occur annually [26,29]. However, the true burden remains uncertain because surveillance relies largely on passive reporting, laboratory confirmation remains incomplete, and many suspected events are not fully investigated or formally classified. Domestic dogs are the principal reservoir and source of human exposures. In Lilongwe, variation in dog population density, movement, and vaccination coverage contributes to spatial heterogeneity in transmission dynamics and surveillance opportunity.

Since 2015, the Lilongwe Society for the Protection and Care of Animals (LSPCA) has coordinated rabies prevention and control activities within the city, including mass dog vaccination, public awareness, and community engagement, establishing collaboration among community, veterinary, public health, and laboratory stakeholders. Up to 2019, rabies surveillance in Lilongwe operated primarily through passive reporting pathways involving communities, the District Health Office (DHO), the District Animal Health and Livestock Development (DAHLD; hereafter referred to as the District Veterinary Office (DVO)), and the Central Veterinary Laboratory (CVL) under DAHLD. Suspected exposures and animal events were reported through routine service channels, with animal investigation and laboratory testing undertaken where feasible. Although these structures supported rabies reporting and confirmation, follow-up and progression from notification to investigation were not systematically coordinated across sectors. The surveillance programme evaluated in this study was implemented within this existing rabies prevention and surveillance system.

### 2.2. Participatory Rabies Surveillance System Design and Implementation

A participatory rabies surveillance programme was implemented in January 2020 through collaboration among the LSPCA, DVO, DHO, CVL, Lilongwe City Council (LCC) and Community Rabies Action Groups (RAGs). This study evaluates surveillance records collected during the first six months of programme implementation (January–June 2020), corresponding to the initial implementation phase of the Community-Based Rabies Surveillance programme.

Surveillance activities were coordinated by four Rabies Project Officers (RPOs) with para-veterinary training who received additional training in rabies epidemiology, bite investigation, and case management. As illustrated in Figure 2, the surveillance system integrated participatory and passive reporting pathways. Participatory surveillance comprised active case-finding, community referrals, and direct reporting via a toll-free hotline (172), whereas passive surveillance consisted of routine reports from the DVO and DHO. Stakeholder coordination was supported through a WhatsApp communication platform and complemented by radio, television, print media, and community sensitisation activities.

**Figure 2.**
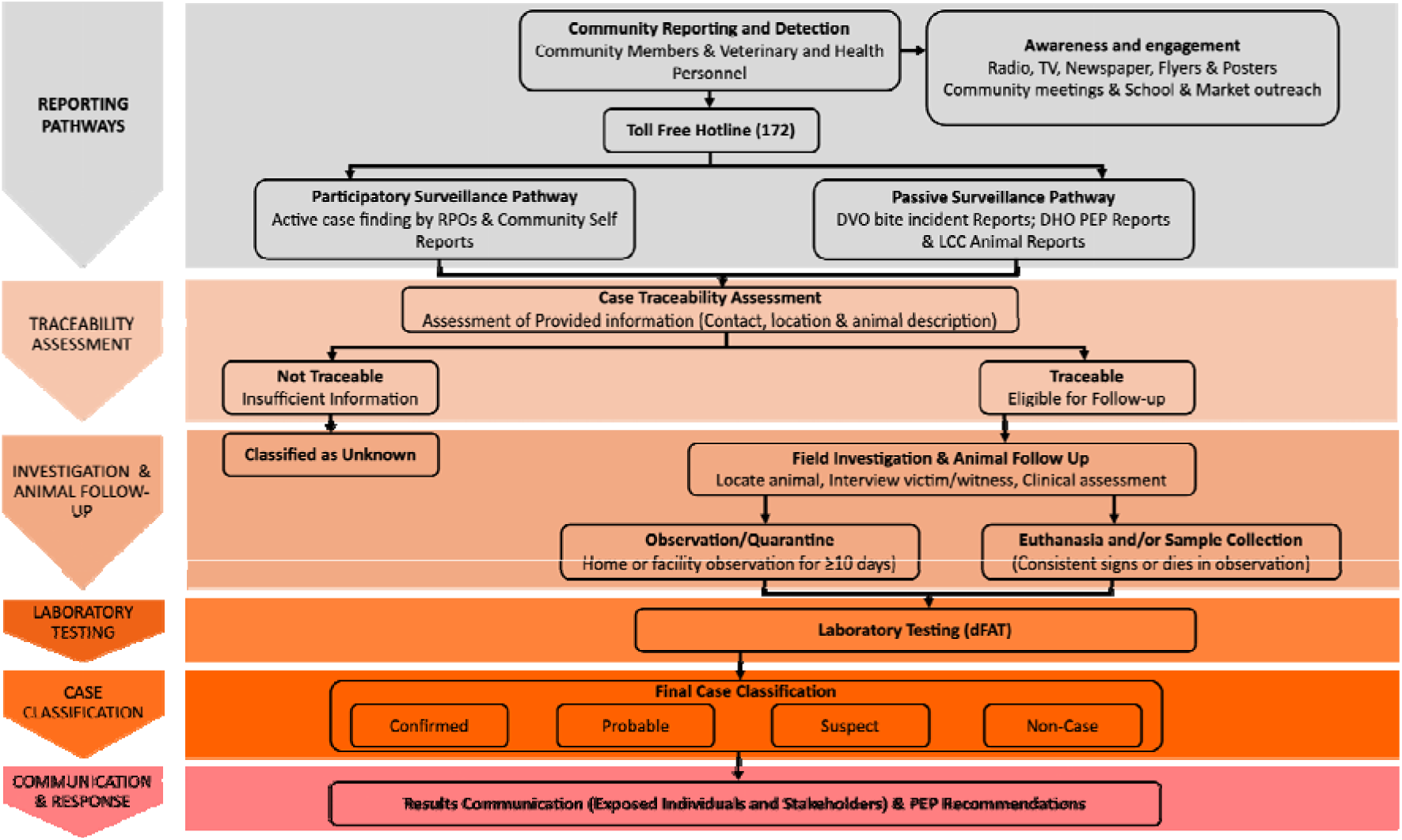
Participatory rabies surveillance and case ascertainment framework. Suspected rabies exposure events entered the surveillance system through participatory and passive reporting pathways. Participatory surveillance included community reporting through a toll-free hotline, community referrals, and active case finding by RPOs, whereas passive surveillance comprised reports originating from DVO, DHO, and LCC. Reported cases underwent traceability assessment, field investigation, animal follow-up, and, where indicated, laboratory testing using the direct fluorescent antibody test (dFAT). Cases were subsequently classified as confirmed, probable, suspect, or non-cases according to standardised surveillance definitions. Results were communicated to relevant stakeholders to support rabies’ risk assessment and PEP decision-making.

Historical DVO bite reports and DHO PEP records were used to identify nine priority areas for targeted deployment of RPOs (Figure 1). Within these areas, RPOs conducted routine patrols, vaccination activities, and active surveillance. However, reporting via the toll-free hotline remained available throughout urban Lilongwe, and all reported events were investigated regardless of location.

### 2.3. Case Investigation and Classification

All reported exposure events were traced through a structured process using available information on the animal, contacts, and location. Tracing was achieved through scheduled telephone interviews and field visits aimed at identifying the implicated animal and assessing its rabies status. Animals without signs suggestive of rabies were placed under quarantine observation for a minimum of 10 days, while those that developed clinical signs consistent with rabies were humanely euthanised in accordance with World Society for the Protection of Animals guidelines [30], with brain tissue collected and submitted to the CVL for diagnosis using the direct fluorescent antibody test. In some instances, where suspected rabid animals had already been killed and buried following community response to aggressive behaviour, carcasses were exhumed to allow laboratory testing. Cases were then classified using standardised surveillance definitions adapted for animal rabies investigation as confirmed, probable, suspect, or non-cases (Supplementary S1) [22], with laboratory findings communicated to relevant stakeholders to inform decisions regarding the initiation or continuation of PEP. Where feasible, photographic documentation of suspected and confirmed cases was collected during field investigations to support verification and communication of findings.

### 2.4. Surveillance Evaluation and Historical Comparison

Surveillance performance was evaluated using a cascade framework tracking progression from reporting to traceability assessment, investigation, and final classification. Attrition between stages was quantified to compare case retention and diagnostic yield across participatory and passive surveillance pathways.

Historical rabies diagnostic records from the Central Veterinary Laboratory were used to establish a pre-surveillance baseline. Comparisons were restricted to corresponding January–June periods from 2015–2019 and the January–June 2020 evaluation period to maintain temporal comparability and minimise seasonal variation in rabies detection.

Data on bite victims linked to investigated cases were reviewed to describe PEP administration according to final rabies classification and to quantify PEP utilisation relative to exposures ultimately classified as rabies.

### 2.5. Data Analysis

Descriptive analyses summarised surveillance outcomes, case classifications, surveillance cascade performance, clinical presentation, and PEP administration. Categorical variables are presented as frequencies and proportions. Surveillance performance was evaluated using a cascade framework, with attrition and retention quantified from reporting through traceability assessment, investigation, and final classification. Differences in case retention between participatory and passive surveillance pathways were assessed using Fisher’s exact test. Diagnostic yield was compared between surveillance pathways by evaluating the odds of identifying confirmed or probable rabies cases among classified events, with effect estimates reported as odds ratios (ORs) and 95% confidence intervals (CIs) derived from Fisher’s exact test. Clinical signs were summarised across rabies classification categories to describe presentation patterns as diagnostic certainty increased. Positive predictive values (PPVs) and exact (Clopper–Pearson) 95% confidence intervals were calculated for individual clinical signs, with confirmed or probable rabies as the outcome. Patterns of PEP administration were described according to the final rabies classification among investigated bite exposures.

Differences in rabies case ascertainment between the historical baseline period (January– June 2015–2019) and the surveillance evaluation period (January–June 2020) were evaluated using negative binomial regression, with log (months under observation) included as an offset term. Effect estimates are reported as incidence rate ratios (IRR) with 95% confidence intervals (CI). All statistical tests were two-sided, and statistical significance was assessed at α = 0.05. Data were analysed in R version 4.4.3 [31].

## 3.0. Results

### 3.1. Characteristics of Reported Rabies Exposure Reports

During the study period, 610 unique suspected rabies events were recorded through the surveillance network. Reports originated through both participatory and routine surveillance pathways. Of these reports, 150 animal events progressed through investigation to final classification and constituted the classified animal-event population for subsequent case-classification analyses. Dogs accounted for nearly all classified animal events (148/150, 98.7%), while cats accounted for the remaining events (2/150, 1.3%).

### 3.2. Surveillance Cascade and Case Retention

Of the 610 reported reports, 239 (39.2%) were considered traceable and eligible for investigation, of which 150 (24.6% of all reports) progressed to final classification (Figure 3). Overall, 371 reports (60.8%) were lost before investigation, and a further 89 (37.2% of traceable reports) did not complete follow-up.

**Figure 3.**
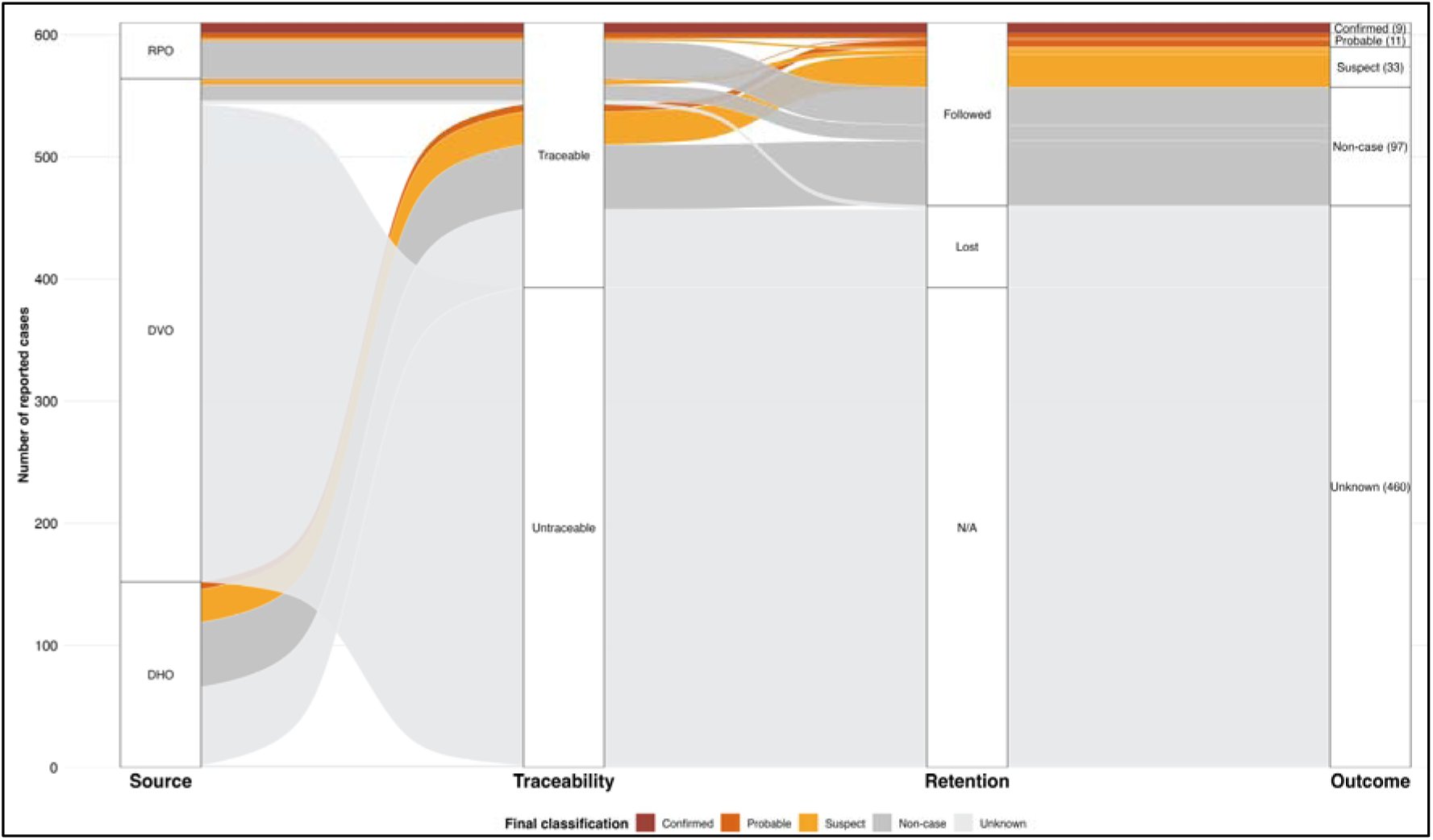
Surveillance progression and investigation outcomes of reported rabies-suspected events by reporting pathways. The surveillance cascade shows progression of reported events from reporting sources through traceability assessment, investigation, and final case classification. Cases lacking sufficient information for investigation or classification were categorised as unknown. Abbreviation: N/A, Not Assessed.

Surveillance performance differed significantly by reporting pathway. All participatory surveillance reports progressed to final classification, whereas only 18.4% of passive reports progressed to classification (46/46 vs 104/564; Fisher’s exact test, p < 0.001). Furthermore, the odds of identifying a confirmed or probable rabies case among classified events were more than five-fold higher under participatory surveillance than passive surveillance (OR = 5.39, 95% CI: 1.82–17.39; p = 0.001). Participatory surveillance identified 13 of 20 (65.0%) confirmed or probable rabies cases and accounted for all laboratory-confirmed cases detected during the evaluation period.

Among the 150 classified events, 97 (64.7%) were classified as non-cases, 33 (22.0%) as suspect, 11 (7.3%) as probable rabies, and 9 (6.0%) as laboratory-confirmed rabies. Together, confirmed and probable cases accounted for 20 (13.3%) of rabies-classified events. Clinical presentation differed across classifications (Supplementary S2). Although biting behaviour was highly prevalent, it had low PPV for confirmed or probable rabies (10.6%, 15/142; 95% CI 6.0–16.8), whereas hypersalivation showed substantially greater PPV (64.3%, 9/14; 95% CI 35.1–87.2).

Detected cases occurred both within and outside areas prioritised for participatory surveillance deployment (Figure 4), with 14 of 20 (70.0%) confirmed or probable cases identified within the nine RPO priority areas.

**Figure 4.**
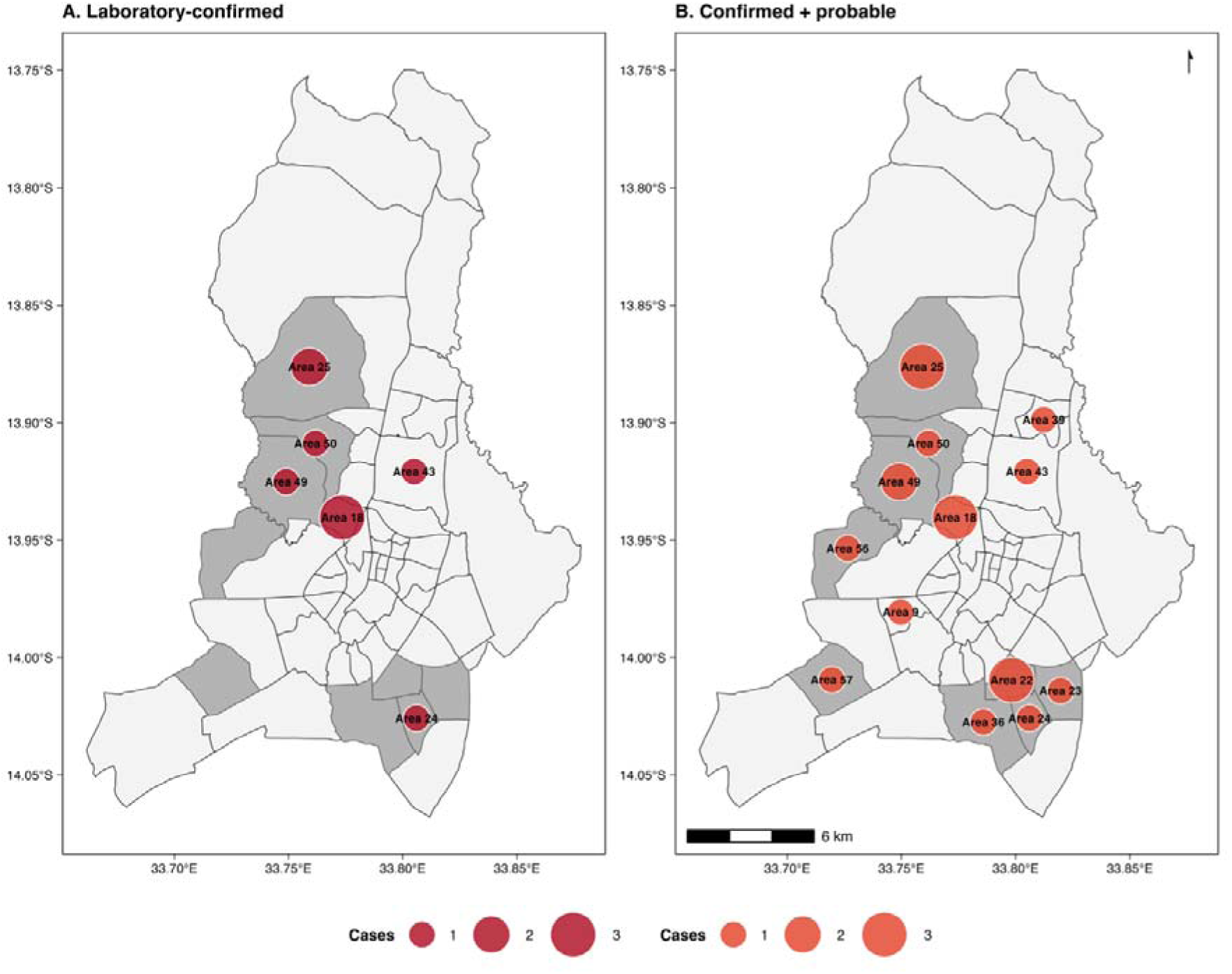
Spatial distribution of rabies cases identified in urban Lilongwe during the surveillance evaluation period. Panel A shows laboratory-confirmed rabies cases, and Panel B shows confirmed and probable rabies cases combined. Grey shading indicates the priority areas where RPOs were deployed, and circle size is proportional to the number of cases identified within each residential area.

Among the 150 classified events, 141 involved human bite exposures and were retained for analyses of exposure characteristics and PEP administration. Dogs accounted for nearly all implicated animals (139/141, 98.6%), whereas cat exposures were uncommon (2/141, 1.4%).

### 3.3. Impact of Participatory Surveillance on Rabies Case Ascertainment

Relative to corresponding January–June periods between 2015 and 2019, participatory surveillance increased rabies case ascertainment during the January–June 2020 evaluation period (Figure 5). Historical surveillance detected a mean of 4.2 laboratory-confirmed rabies cases per the January–June period, whereas the surveillance programme identified 20 rabies-classified cases comprising 9 laboratory-confirmed and 11 probable cases. Restricting the comparison to laboratory-confirmed detections alone yielded a 2.4-fold increase in rabies detection. When probable cases were included, overall rabies case ascertainment increased to 4.8-fold (IRR 4.76, 95% CI 1.32–22.87; p=0.027).

**Figure 5.**
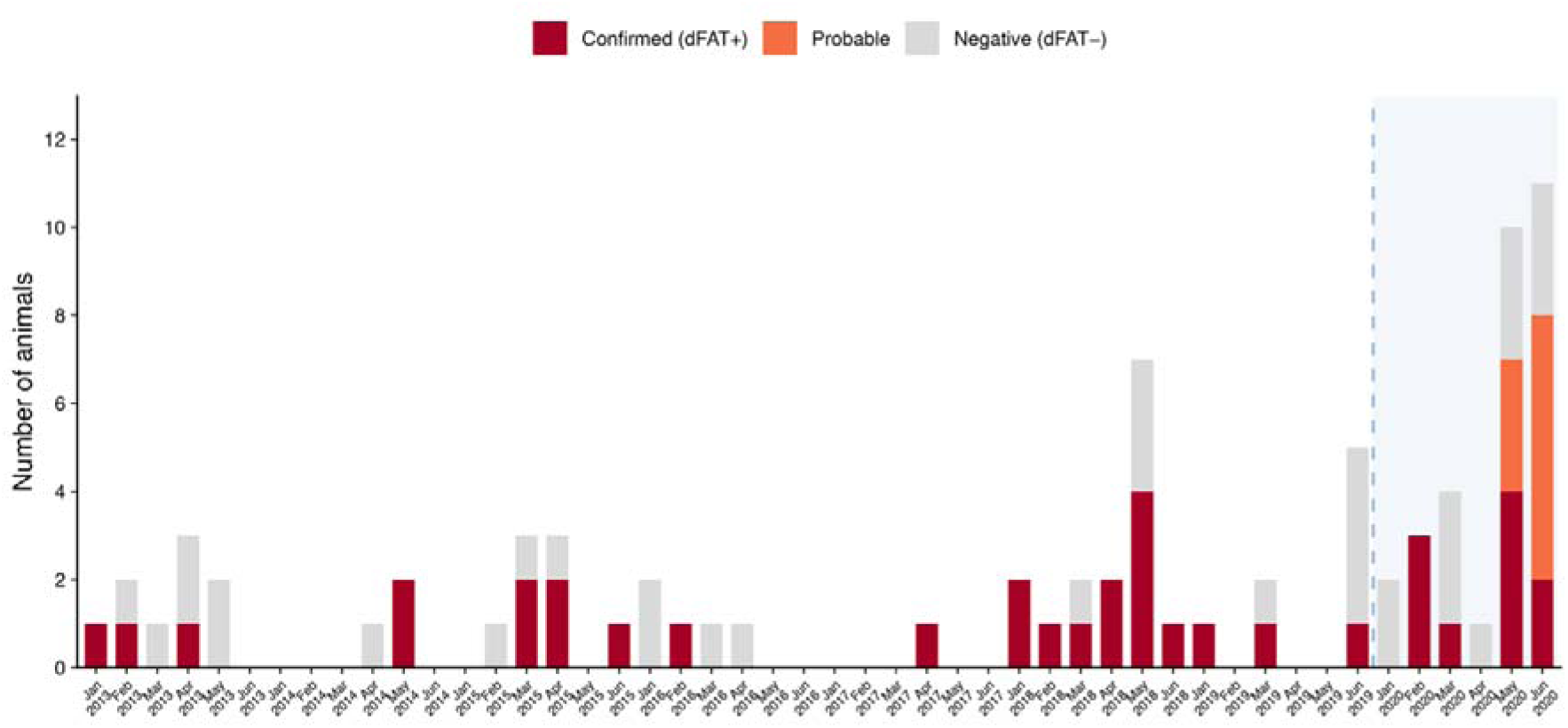
Comparison of rabies case ascertainment during corresponding January–June periods before and during participatory surveillance implementation in Lilongwe. Bars represent monthly rabies investigation outcomes. Historical observations (2015–2019) include CVL laboratory outcomes only (dFAT-confirmed and dFAT-negative cases), whereas the January–June 2020 surveillance period additionally includes probable rabies cases identified through participatory surveillance. The shaded region and dashed line indicate the participatory surveillance period.

### 3.4. Human Bites and Corresponding PEP Issuance

Among the 141 bite victims linked to investigated exposure events, 92 (65.2%) were exposed to animals ultimately classified as non-cases, 33 (23.4%) to suspect cases, 9 (6.4%) to probable rabies cases, and 7 (5.0%) to laboratory-confirmed rabies cases.

Overall, 136 of 141 bite victims (96.5%) received rabies PEP. PEP administration was universal across exposures classified as suspect, probable, or confirmed rabies and remained high among exposures classified as non-cases: 87/92 (94.6%; 95% CI 87.8–98.2%). Of the 136 individuals receiving PEP, only 16 (11.8%) were linked to confirmed or probable rabies exposures, whereas 87 (64.0%) were associated with exposures ultimately classified as non-cases (Figure 6). This corresponds to a PEP-to-rabies exposure ratio of 8.5:1, indicating that approximately nine individuals initiated PEP for every exposure classified as confirmed or probable rabies.

**Figure 6.**
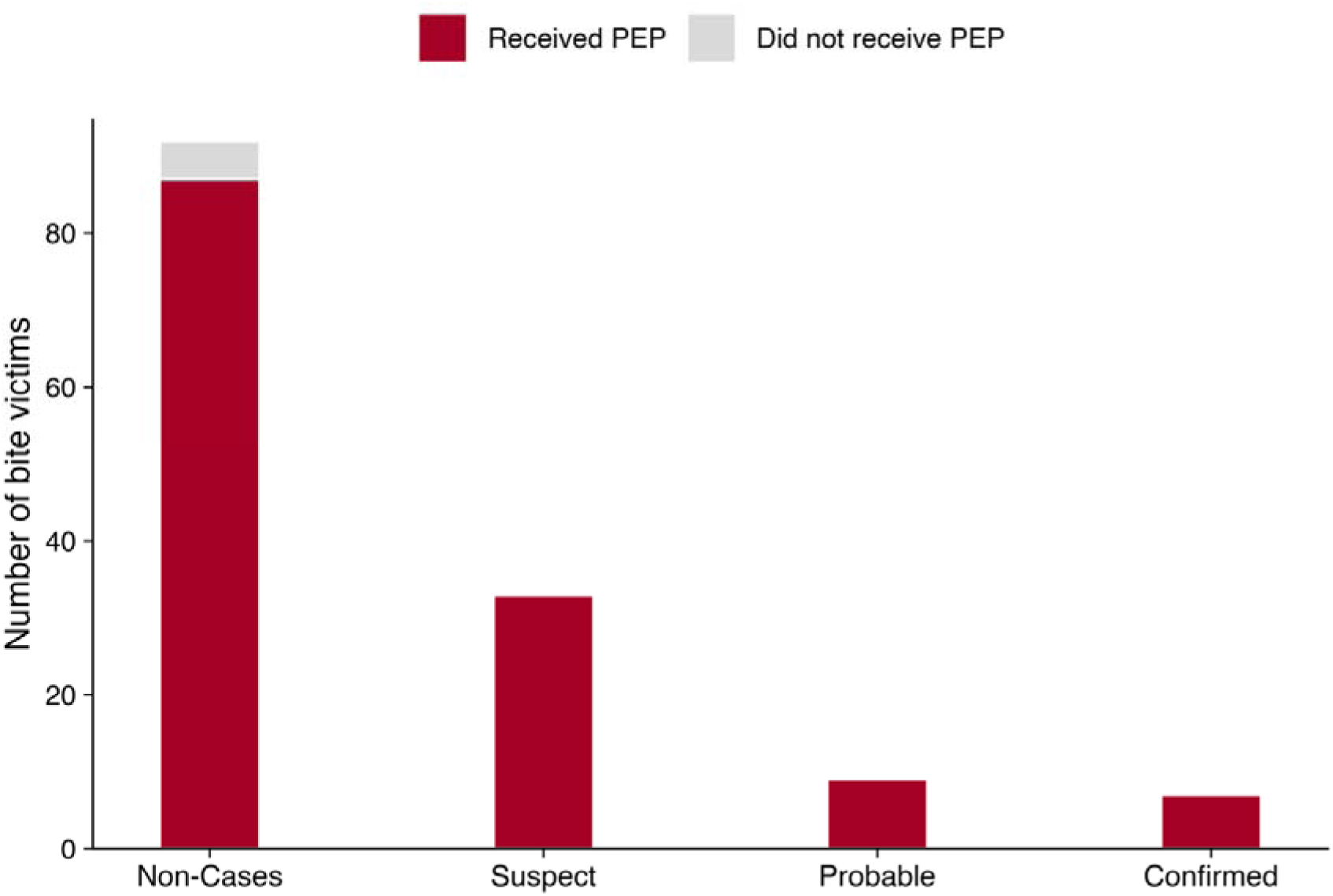
Distribution of rabies PEP administration across final rabies classifications among investigated bite exposures in Lilongwe, Malawi. Bars represent the absolute number of bite victims stratified by the final classification of the implicated animal and receipt of PEP.

## 4.0 Discussion

This study demonstrates that participatory rabies surveillance strengthened rabies case detection in urban Lilongwe and provides insight into how these gains occurred across the surveillance cascade. Relative to corresponding historical surveillance periods, implementation of participatory surveillance was associated with a nearly fivefold increase in rabies case ascertainment and resulted in the identification of all laboratory-confirmed rabies cases detected during the evaluation period. These findings are consistent with previous studies showing that strengthened and community-linked surveillance approaches substantially increase rabies detection in endemic settings [12,21,22,32,33]. Our findings suggest that improvements in ascertainment may arise not simply from generating more reports but from improving continuity of surveillance processes from initial notification through investigation and diagnostic resolution.

Our findings suggest that surveillance performance is determined not only by the number of suspected events reported but also by where cases are lost as they progress through the surveillance pathway. The surveillance cascade identified substantial losses after cases entered the surveillance system, with the greatest attrition occurring before investigation and final classification. Many reports lacked sufficient contact or location information to enable tracing of the reporter or animal, while others did not progress through follow-up and diagnostic resolution. These findings suggest that rabies under-ascertainment arises not only from missed case reporting but also from cumulative operational losses during traceability, investigation, and final classification. Similar operational constraints, including delayed reporting, incomplete traceability [2,34], fragmented information systems [34–36], and inconsistent follow-up, have been identified as persistent barriers to effective rabies surveillance and control in endemic settings. In contrast, the participatory pathway appeared to reduce losses during traceability assessment and investigation completion, improving progression of suspected rabies cases from notification to diagnostic resolution and final classification. In this study, participatory surveillance was associated with higher retention across the surveillance cascade, with all participatory reports progressing to final classification compared with 18.4% of passive reports, and was associated with higher diagnostic yield among classified events. These findings suggest that surveillance performance depends not only on the volume of incoming reports but also on the ability of surveillance systems to retain reported events through to diagnostic resolution. Participatory surveillance may therefore strengthen routine surveillance by increasing the conversion of reported events into actionable surveillance outcomes through community-linked reporting, active follow-up, and coordination across veterinary, laboratory, and public health sectors. This supports the integration of participatory approaches within existing One Health surveillance systems [9,37,38]. Furthermore, the detection of cases outside RPO priority areas also underscores the need for surveillance systems to preserve follow-up capacity beyond initially prioritised locations.

The clinical presentation patterns highlight the importance of structured follow-up after animal bite reports. Bite events are important entry points into rabies surveillance and appropriate triggers for precautionary public health action, but they provide limited information on rabies status unless followed by systematic animal investigation. In this study, the value of clinical and epidemiological assessment was not to replace laboratory confirmation, but to support more consistent classification of reported events. Strengthening this process may improve the quality of surveillance information available for exposure risk assessment and coordination between veterinary and public health services.

Beyond case detection, our findings show how surveillance performance can shape public health decision-making and resource allocation at the point of care. Although PEP initiation was nearly universal among investigated bite victims, only 11.8% of PEP recipients were ultimately linked to confirmed or probable rabies exposures, corresponding to a PEP-to-rabies exposure ratio of 8.5:1. This pattern should not be interpreted as inappropriate treatment, as precautionary PEP initiation is consistent with WHO guidance when rabies exposure cannot be excluded [39], and similar patterns have been reported where animal investigations are delayed or incomplete [40–42]. Rather, it illustrates how uncertainty within the animal surveillance pathway can propagate into human clinical decision-making.

Our findings help locate this uncertainty within a measurable surveillance bottleneck. When reported animals are not traced, investigated, or classified in time, clinicians must initiate PEP without animal outcome information. Losses before investigation may therefore contribute to precautionary PEP administration and reduce the efficiency of limited PEP resources. This has practical implications in settings where PEP remains expensive, supply-constrained, or unevenly accessible [43,44], because avoidable treatment courses carry both direct costs and opportunity costs for individuals at genuine risk [33,34,45–48].

Strengthening traceability assessment, timely animal investigation [33,45], rapid communication of animal outcomes to clinicians [21,46], and veterinary-public health coordination [34], could allow PEP decisions to be revised as new information becomes available. Framed in this way, participatory surveillance is not merely a case-detection tool, but a mechanism for reducing diagnostic uncertainty and supporting safer, more targeted use of scarce PEP resources.

## 5.0 Limitations

Several limitations should be considered. The evaluated surveillance activities occurred during the COVID-19 pandemic, and associated restrictions may have differentially affected surveillance pathways by altering mobility, healthcare-seeking behaviour, and institutional operations. These shifts may have influenced case reporting and follow-up, meaning that some observed differences in retention and case ascertainment may partly reflect the pandemic’s contextual effects rather than surveillance design alone. In addition, clinical signs were not uniformly observed across investigations, and some animals were euthanised before complete clinical assessment. Historical comparisons relied on laboratory surveillance records and may not fully account for temporal changes in reporting practices.

## 6.0 Conclusion

This study demonstrates that rabies cases are lost not only before they are reported but also after entering surveillance systems, particularly during traceability assessment, investigation, and final classification. Participatory surveillance reduced these losses by improving progression through the surveillance cascade, resulting in greater case ascertainment than routine passive surveillance. Beyond disease detection, improved surveillance may support more informed assessment of exposure risk, strengthen coordination between the animal and human health sectors, and enhance the targeting of interventions, including PEP. These findings support integration of participatory approaches within existing surveillance structures as a practical strategy for strengthening rabies surveillance and response.

## Data Availability

The data underlying this study are owned by the Government of Malawi and are not publicly available. Requests for access should be directed to the Department of Animal Health and Livestock Development (DAHLD) and the Lilongwe District Health Office (DHO with involvement of the Lilongwe Society for the Protection and Care of Animals (LSPCA) where applicable, subject to relevant approvals and data-sharing requirements.

## 7.0 Supplementary

Supplementary S1: Case definitions used for classification of suspected rabies events

Supplementary S2: Distribution and positive predictive values of clinical signs among investigated animal rabies reports

## List of abbreviations

CBRS: Community-Based Rabies Surveillance
CI: Confidence Interval
CVL: Central Veterinary Laboratory
DAHLD: District Animal Health and Livestock Development
DHO: District Health Office
DVO: District Veterinary Office
IBCM: Integrated Bite Case Management
LCC: Lilongwe City Council
LSPCA: Lilongwe Society for the Protection and Care of Animals
OR: Odds Ratio
PEP: Post-Exposure Prophylaxis
RAG: Rabies Action Group
RPO: Rabies Project Officer
WHO: World Health Organization

## 8.0 Ethics Approval and Consent to Participate

Ethical Approval for the study was provided by the Department of Animal Health and Livestock Development Research Ethics Committee, reference number DAHLD/AHC/12/2019/1. The requirement for individual informed consent was waived by the Department of Animal Health and Livestock Development Research Ethics Committee because the analysis used routinely collected programme data. All records used for analysis were anonymised and contained no personal identifiers.

## 9.0 Consent for Publication

Not applicable.

## 11.0 Competing Interests

The authors declare that they have no known competing financial interests or personal relationships that could have appeared to influence the work reported in this paper.

## 12.0 Funding

This research was generously supported by the Lilongwe University of Agriculture and Natural Resources (LUANAR), the Lilongwe Society for the Protection and Care of Animals, and the Martyn Edelsten (of the University of Edinburgh) Research and Internship Grant Fund.

## 13.0 Authors’ Contributions

PIM: Writing – review & editing, Writing – original draft, Conceptualisation, Visualisation, Validation, Software, Methodology, Investigation, Formal analysis, Funding acquisition, Data curation. PKK: Validation, Writing – review & editing. EC: Writing – review & editing, Resources, Project administration, Investigation, Conceptualisation. TR: Writing – review & editing, Conceptualisation, Project administration. MN: Writing – review & editing, Supervision, Conceptualisation, Project administration. GC: Writing – review & editing, Project administration. KMP: Investigation, Writing – review & editing. DL: Investigation, Writing – review & editing. PD: Investigation, Writing – review & editing. Loveness Kamanga: Investigation, Writing – review & editing. MM: Investigation, Writing – review & editing. MT: Writing – review & editing, Validation, Supervision, Methodology, Funding acquisition, Conceptualisation. TFK: Writing – review & editing, Validation, Supervision.

## 14.0 Acknowledgements

We would like to thank Lieza Swenen, Kondwani Chiumya, Julia Pascarella, Dr Maike Müller, and the entire partner teams for their expertise and assistance throughout all aspects of this work.

## Notes

### Competing Interest Statement

The authors have declared no competing interest.

### Summary of Updates

This version reflects revisions to the analysis, interpretation, and presentation of the surveillance pathways. The manuscript provides a clearer account of case ascertainment and attrition across the surveillance cascade, including where suspected cases are retained or lost between reporting, tracing, investigation, and final classification. The analysis and interpretation of post-exposure prophylaxis use have also been updated. Figures, tables, terminology, and the Discussion have been revised for consistency with these updates. The author list has also been updated to include additional contributors who made substantive intellectual and technical contributions to the revised manuscript.

