## Supplementary S1 for "Where Are Rabies Cases Lost Along the Surveillance Pathway? A Comparative Evaluation of Participatory and Passive Rabies Surveillance in Urban Lilongwe, Malawi"

**Supplementary S1** Operational case definitions used for classification of suspected rabies events during surveillance evaluation. Final classification incorporated available laboratory, clinical, and epidemiological evidence.

| Classification | Case Definition |
|---|---|
| Confirmed | Diagnostic confirmation of rabies virus infection by direct fluorescent antibody test (dFAT). |
| Probable | Animals without laboratory confirmation but with epidemiological or clinical evidence consistent with rabies, including: (i) animals that died during observation following suspected rabies exposure; (ii) animals that developed compatible clinical signs and subsequently died; or (iii) animals epidemiologically linked to a confirmed, probable, or suspect rabid animal. |
| Suspect | Animals reported as potentially rabid but lacking sufficient clinical, laboratory, or epidemiological evidence for classification as probable or confirmed, including animals that could not be fully assessed or had inconclusive investigations. |
| Non-case | Animals that remained healthy following the quarantine period or tested negative for rabies by dFAT. |
