## Supplementary S2 for "Where Are Rabies Cases Lost Along the Surveillance Pathway? A Comparative Evaluation of Participatory and Passive Rabies Surveillance in Urban Lilongwe, Malawi"

**Supplementary S2** Distribution and positive predictive values of clinical signs among investigated animal rabies reports by final case classification.

| **Signs** | **Non-Case** (n=97) | **Suspect** (n=33) | **Probable** (n=11) | **Confirmed** (n=9) | **Confirmed or Probable cases with Sign,** n | **PPV (95% CI)** |
|---|---|---|---|---|---|---|
| Paresis/paralysis | 0 (0.0%) | 0 (0.0%) | 2 (18.2%) | 1 (11.1%) | 3 | 100.0% (29.2–100.0)* |
| Aggression | 11 (11.3%) | 1 (3.0%) | 5 (45.5%) | 4 (44.4%) | 9 | 42.9% (21.8–66.0) |
| Abnormal vocalisation | 5 (5.2%) | 0 (0.0%) | 2 (18.2%) | 2 (22.2%) | 4 | 44.4% (13.7–78.8) |
| Biting | 94 (96.9%) | 33 (100.0%) | 9 (81.8%) | 6 (66.7%) | 15 | 10.6% (6.0–16.8) |
| Hypersalivation | 5 (5.2%) | 0 (0.0%) | 5 (45.5%) | 4 (44.4%) | 9 | 64.3% (35.1–87.2) |
| Lethargy | 0 (0.0%) | 0 (0.0%) | 0 (0.0%) | 1 (11.1%) | 1 | 100.0% (2.5–100.0)* |
| Restlessness | 4 (4.1%) | 0 (0.0%) | 3 (27.3%) | 1 (11.1%) | 4 | 50.0% (15.7–84.3) |
| Anorexia | 4 (4.1%) | 0 (0.0%) | 1 (9.1%) | 3 (33.3%) | 4 | 50.0% (15.7–84.3) |
| Hydrophobia | 0 (0.0%) | 0 (0.0%) | 0 (0.0%) | 1 (11.1%) | 1 | 100.0% (2.5–100.0)* |
| Automobile accident | 2 (2.1%) | 0 (0.0%) | 0 (0.0%) | 0 (0.0%) | 0 | 0.0% (0.0–84.2)* |

Abbreviations: CI, confidence interval; PPV, positive predictive value.

* Estimates based on fewer than five observations should be interpreted cautiously.
